# Heart size disparity drives sex-specific response to cardiac resynchronization therapy: a post-hoc analysis of the MORE-MPP CRT trial

**DOI:** 10.1101/2023.12.05.23299532

**Authors:** Nadeev Wijesuriya, Vishal Mehta, Felicity De Vere, Sandra Howell, Steven A Niederer, Haran Burri, Johannes Sperzel, Leonardo Calo, Bernard Thibault, Wenjiao Lin, Kwangdeok Lee, Andrea Grammatico, Niraj Varma, Marianne Gwechenberger, Christophe Leclercq, Christopher A Rinaldi

## Abstract

**Background:** Studies have reported that female sex predicts superior cardiac resynchronization therapy (CRT) response. One theory is that this association is related to smaller female heart size, thus increased “relative dyssynchrony” at given QRS durations (QRSd).

**Objective:** To investigate the mechanisms of sex-specific CRT response relating to heart size, relative dyssynchrony, cardiomyopathy type, QRS morphology, and other patient characteristics.

**Methods:** A post-hoc analysis of the MORE-CRT MPP trial (n=3739, 28% female), with a sub-group analysis of patients with non-ischaemic cardiomyopathy (NICM) and left bundle branch block (LBBB) (n=1308, 41% female) to control for confounding characteristics. A multivariable analysis examined predictors of response to 6 months of conventional CRT, including sex and relative dyssynchrony, measured by QRSd/LVEDV (left ventricular end-diastolic volume).

**Results:** Females had a higher CRT response rate than males (70.1% vs. 56.8%, p<0.0001). *Subgroup analysis:* Regression analysis of the NICM LBBB subgroup identified QRSd/LVEDV, but not sex, as a modifier of CRT response (p<0.0039). QRSd/LVEDV was significantly higher in females (0.919) versus males (0.708, p<0.001). CRT response was 78% for female patients with QRSd/LVEDV>median value, compared to 68% < median value (p=0.012). Association between CRT response and QRSd/LVEDV was strongest at QRSd<150ms.

**Conclusions:** In the NICM LBBB population, increased relative dyssynchrony in females, who have smaller heart sizes than their male counterparts, is a driver of sex-specific CRT response, particularly at QRSd <150ms. Females may benefit from CRT at a QRSd <130ms, opening the debate on whether sex-specific QRSd cut-offs or QRS/LVEDV measurement should be incorporated into clinical guidelines.

## Introduction

Cardiac Resynchronization Therapy (CRT) is a hallmark treatment for patients with dyssynchronous heart failure.(1) However, despite its widespread success and uptake, 30% of patients fail to derive benefit.(2,3) These “CRT non-responders” have amongst the poorest long-term outcomes of any subgroup in the heart failure population.(4) As such, there is significant interest in examining factors which modulate CRT response.

It has been well established that certain conditions are associated with a poorer clinical or LV remodelling responses to CRT, for example, ischaemic cardiomyopathy, atrial fibrillation (AF) and non-left bundle branch block (LBBB) QRS morphology.(5) These associations are unsurprising from a physiological perspective. An area which remains poorly understood is the association of male or female sex in CRT response. Although some studies report no sex differences, for example CARE-HF(6) most have reported that female sex is predictive of superior clinical and echocardiographic response.(7–12) In a meta-analysis of 149,259 patients, Yin et al. observed a lower all-cause mortality in females than males post-CRT (odds ratio[OR] 0.50, 95% confidence interval [CI] 0.36 to 0.70).(13) Females also exhibited statistically significant improvement in left ventricular ejection fraction (LVEF) and decrement of left ventricular end-diastolic diameter (LVEDD) when compared with males.

The mechanisms underpinning this association remain unclear. One theory is that the higher female response rate is related to a higher frequency of LBBB and non-ischaemic cardiomyopathy (NICM) phenotypes. Determining this accurately requires examination of large sample sizes, especially as women represent only 20-30% of the population in CRT trials.(14)

Sex-related CRT response disparity may also be explained by differences in cardiac size,(15) that is, that females are relatively smaller than males, and therefore have a greater degree of dyssynchrony at a given QRSd. This is supported by studies that report that females exhibit an improved CRT response compared to males in cohorts with a QRSd <150ms.(16–18). It is suggested that sex-specific differences in QRS-response relationship are unexplained by the application of strict LBBB criteria or by body surface area, but resolved by QRSd “normalization” for heart size using LV mass or LV end diastolic volume (LVEDV).(19,20) These metrics of “relative dyssynchrony” such as QRSd/LVEDV are not routinely measured in clinical practice, nor are they frequently reported in larger CRT trials. As such, this area has to date only been examined in single-center cohorts with small sample sizes. A meta-analysis of 3496 patients identified height and QRS duration, but not sex, as independent predictor for the composite outcome of first hospitalisation for heart failure and all-cause mortality, however, QRSd/LVEDV was not used in this analysis.(21)

In this study, we aimed to determine the association between sex, LV size and relative dyssynchrony in a large cohort of CRT recipients. We tested this by performing a sub-analysis of patients recruited to the MORE-CRT MPP trial.

## Methods

MORE-CRT MPP is a prospective, randomized, multicenter study.(22,23) All patients initially receive conventional biventricular (BiV) CRT for 6 months. At this stage, echocardiographic non-responders are randomized to either continued conventional BiV pacing, or multi-point pacing. CRT response was defined as a reduction in LV end systolic volume (LVESV) >15% analysed by an independent core lab. We evaluated data following the first 6 months of conventional BiV CRT; a post-hoc multivariable analysis examined predictors of this reverse remodelling response (Supplement Figure 1). In particular we assessed the predicted probability of CRT response according to sex and QRSd/LVEDV. A subgroup analysis of the NICM LBBB population was performed to test the effect without confounding from significant variables such as QRS morphology and cardiomyopathy phenotype.

### Study participants

The study enrolled eligible patients with a standard CRT indication after obtaining written informed consent. The device was programmed BiV pacing in those patients who had the quadripolar CRT system successfully implanted (Quartet LV lead with a Quadra CRT device, Abbott, Sylmar, CA, USA) with programming of the LV pacing vector, A-V delay, and V-V delay settings at the implanting physician’s discretion. During the first 6 months following implant, all subjects received standard BiV pacing.

### Echocardiographic assessment

Trained and qualified site personnel performed echocardiographic recordings, with analysis performed by an independent Echocardiography Core Laboratory. The core laboratory averaged the LV volumes measured in the apical two-chamber view and four-chamber view and measured LVEF. All measurements were made at baseline and at 6 months.

### Statistics

Summary statistics were used for baseline characteristics. Continuous variables were summarized by means and SD. Two-sample student’s t-test or Wilcoxon rank sum test was used to test the difference between 2 groups depending on the normality of the data. Frequencies and percentages summarized categorical variables. Chi-square test or Fisher exact test was used to test the difference between groups.

We performed univariable models using the following baseline variables: age, AF, chronic obstructive pulmonary disease (COPD), diabetes, hypercholesterolaemia, hypertension, LVEDV, LVEF, New York Heart Association (NYHA) Class I/II versus NYHA III/IV, renal disease, ischaemic versus NICM, LBBB versus non-LBBB, QRSd, QRSd/LVEDV (“normalised QRS) and sex. Multivariable models were conducted using stepwise selections. The criteria for baseline variables entering and staying into model was α=0.25 and α=0.05, respectively. Age and gender were forced into the model. Given that logistic regression methods relies upon the assumption of linearity between CRT response and the predicting independent variables (patients’ characteristics) we also evaluated the probability of CRT response as a function of QRSd (divided in 5 subgroups) and as a function of QRSd normalized by dividing it by LVEDV (divided in 10 deciles) to account for heart size differences between females and males. The predicted probability of CRT response as a function of QRSd and normalised QRSd was derived from multivariable logistic regression models. Analyses were conducted using SAS^®^ 9.4.

## Results

### Baseline Characteristics

The analysis was performed on 3739 patients (1051, 28% female). Patient baseline characteristics are shown in Tables 1 and 2. In the whole population (Table 1), the cardiac size in females was significantly smaller compared to males (176±60 ml vs. 228 ±77 ml, p<0.0001). Females had a significantly shorter QRSd than males (153±22 ms vs.158±26 ms, p<0.0001). Similar results on cardiac size and QRSd were observed in the non-ischemic LBBB subgroup (Table 2),

**Table 1.** Baseline patient characteristics for the whole population. LBBB = Left Bundle Branch Block; COPD = Chronic Obstructive Pulmonary Disease; ACE = Angiotensin-converting enzyme; ARB = Angiotensin II receptor blocker; *Wilcoxon rank sum test, ^†^Pearson chi-square test

| Demographic Variable | All Subjects<br>(N=3,739) | Female<br>Subjects<br>(N=1,051) | Male Subjects<br>(N=2,688) | P value |
| --- | --- | --- | --- | --- |
| **Age (Years)** |  |  |  |  |
| Mean ± SD | 68 ± 11 | 68 ± 11 | 68 ± 11 | 0.3454* |
| **NYHA Class at Enrolment (%)** |  |  |  |  |
| Class II | 51.2 | 45.3 | 53.5 | <0.0001† |
| Class III | 46.6 | 52.1 | 44.5 |  |
| Class IV | 1.9 | 2.4 | 1.7 |  |
| **QRS Duration (ms)** |  |  |  |  |
| Mean ± SD (n) | 156 ± 25 | 153 ± 22 | 158 ± 26 | <0.0001* |
| **QRS Morphology (%)** |  |  |  |  |
| LBBB | 70.2 | 79.5 | 66.4 | <0.0001† |
| Non-LBBB | 29.8 | 20.5 | 33.6 |  |
| **Cardiomyopathy Etiology (%)** |  |  |  |  |
| Ischemic | 41.0 | 22.0 | 48.4 | <0.0001† |
| Non-Ischemic | 59.0 | 78.0 | 51.6 |  |
| **LVESV (ml)** |  |  |  |  |
| Mean ± SD | 160 ± 65 | 131 ± 52 | 171 ± 66 | <0.0001* |
| **LVEDV (ml)** |  |  |  |  |
| Mean ± SD | 214 ± 76 | 176 ± 60 | 228 ± 77 | <0.0001* |
| **LVEF (%)** |  |  |  |  |
| Mean ± SD | 26 ± 7 | 27 ± 7 | 26 ± 7 | 0.0965* |
| **Device Type (%)** |  |  |  |  |
| CRT-P | 12.2 | 16.9 | 10.4 | <0.0001† |
| CRT-D | 87.8 | 83.1 | 89.6 |  |
| **Medical History (%)** |  |  |  |  |
| Hypertension | 62.3 | 59.0 | 63.6 | 0.0087† |
| Hypercholesterolemia | 40.1 | 34.6 | 42.2 | <0.0001† |
| Diabetes | 33.4 | 30.4 | 34.6 | 0.0126† |
| COPD | 10.6 | 8.5 | 11.5 | 0.0076† |
| Renal Disease | 15.0 | 11.4 | 16.4 | 0.0001† |
| **Medical Treatment (%)** |  |  |  |  |
| Diuretics | 77.4 | 79.5 | 76.5 | 0.0474† |
| ACE inhibitor or ARB | 89.3 | 89.4 | 89.2 | 0.8400† |
| β-Blocker | 89.1 | 89.2 | 89.1 | 0.8697† |
| Aldosterone antagonist | 38.9 | 38.5 | 39.1 | 0.7660† |
| Anticoagulant | 26.9 | 21.4 | 29.1 | <0.0001† |
| Calcium Channel Blockers | 7.9 | 6.2 | 8.6 | 0.0156† |
| Nitrates | 7.4 | 5.6 | 8.1 | 0.0097† |

**Table 2.** Baseline patient characteristics for non-ischemic LBBB patients. COPD = Chronic Obstructive Pulmonary Disease; ACE = Angiotensin-converting enzyme; ARB = Angiotensin II receptor blocker; *Wilcoxon rank sum test, ^†^Pearson chi-square test

| Demographic Variable | All Subjects<br>(N=1,308) | Female<br>Subjects<br>(N=538) | Male Subjects<br>(N=770) | P value |
| --- | --- | --- | --- | --- |
| **Age (Years)** |  |  |  |  |
| Mean ± SD | 66 ± 11 | 67 ± 10 | 65 ± 11 | 0.0039* |
| **NYHA Class at Enrolment (%)** |  |  |  |  |
| Class II | 55.4 | 45.5 | 62.2 | <0.0001† |
| Class III | 42.2 | 51.5 | 35.7 |  |
| Class IV | 2.1 | 2.6 | 1.8 |  |
| **QRS Duration (ms)** |  |  |  |  |
| Mean ± SD (n) | 161 ± 19 | 157 ± 18 | 165 ± 20 | <0.0001* |
| **LVESV (ml)** |  |  |  |  |
| Mean ± SD | 162 ± 70 | 135 ± 54 | 181 ± 73 | <0.0001* |
| **LVEDV (ml)** |  |  |  |  |
| Mean ± SD | 216 ± 82 | 180 ± 64 | 241 ± 85 | <0.0001* |
| **LVEF (%)** |  |  |  |  |
| Mean ± SD | 26 ± 7 | 26 ± 7 | 26 ± 7 | 0.3278* |
| **Device Type (%)** |  |  |  |  |
| CRT-P | 11.3 | 13.9 | 9.5 | 0.0122† |
| CRT-D | 88.7 | 86.1 | 90.5 |  |
| **Medical History (%)** |  |  |  |  |
| Hypertension | 56.4 | 55.0 | 57.4 | 0.3922† |
| Hypercholesterolemia | 33.1 | 29.9 | 35.3 | 0.0412† |
| Diabetes | 28.4 | 29.4 | 27.8 | 0.5342† |
| COPD | 9.7 | 8.9 | 10.3 | 0.4213† |
| Renal Disease | 9.2 | 9.1 | 9.2 | 0.9445† |
| **Medical Treatment (%)** |  |  |  |  |
| Diuretics | 76.6 | 78.8 | 75.1 | 0.1154† |
| ACE inhibitor or ARB | 91.5 | 90.7 | 92.1 | 0.3811† |
| β-Blocker | 91.1 | 90.3 | 91.6 | 0.4454† |
| Aldosterone antagonist | 40.0 | 38.3 | 41.2 | 0.2956† |
| Anticoagulant | 21.0 | 17.7 | 23.4 | 0.0125† |
| Calcium Channel Blockers | 5.0 | 5.2 | 4.9 | 0.8266† |
| Nitrates | 3.7 | 3.5 | 3.8 | 0.8242† |

### CRT response in the whole study cohort

CRT response by sex is outlined in Table 3. Female patients had a higher CRT response rate than males in the total population (70.1% vs. 56.8%, p<0.0001). The results of the logistic regression model (Table 4) show that LBBB, wide QRSd, and female sex are significant independent predictors for improved response while history of AF, ischemic aetiology, large heart (LVEDV), and history of renal disease are significant independent predictors for reduced CRT response. When QRSd/LVEDV ratio instead of QRSd was included in a separate model, QRSd/LVEDV was not a significant independent predictor of response (OR 1.21 95% CI 0.89-1.63, p=0.22).

**Table 3.** CRT response by sex.

| Population | Gender | 6M Responder Rate(n/N) | p-value |
| --- | --- | --- | --- |
| Whole Population | F | 70.12% (737/1051) | <.0001 |
|  | M | 56.85% (1528/2688) |  |
| LBBB Patients | F | 71.07% (479/674) | <.0001 |
|  | M | 62.06% (854/1376) |  |
| Non-ischemic LBBB Patients | F | 73.05% (393/538) | 0.2508 |
|  | M | 70.13% (540/770) |  |
| Non-ischemic LBBB and QRSd<150ms | F | 69.71% (122/175) | 0.0157 |
|  | M | 56.86% (87/153) |  |
| Non-LBBB Patients | F | 62.64% (109/174) | <.0001 |
|  | M | 40.29% (280/695) |  |

**Table 4.** Logistic regression results (whole population)

| Parameter | Univariate |  |  | Multivariate (2866 patients) |  |
| --- | --- | --- | --- | --- | --- |
|  | Parameter Estimate<br>[95% CI] | p<br>value | Sample<br>Size | Parameter Estimate<br>[95% CI] | p value |
| Age | 0.998 [0.992, 1.004] | 0.5685 | 3,738 | 1.002 [0.994, 1.010] | 0.6972 |
| Atrial fibrillation | 0.679 [0.565, 0.815] | <.0001 | 3,739 | 0.785 [0.627, 0.982] | 0.0339 |
| COPD | 1.018 [0.822, 1.260] | 0.8702 | 3,739 |  |  |
| Diabetes | 0.770 [0.671, 0.884] | 0.0002 | 3,739 |  |  |
| Hypercholesterolemia | 0.813 [0.712, 0.929] | 0.0024 | 3,739 |  |  |
| Hypertension | 1.047 [0.914, 1.198] | 0.5096 | 3,739 |  |  |
| Ischemic vs<br>Non-Ischemic | 0.502 [0.439, 0.574] | <.0001 | 3,739 | 0.550 [0.466, 0.648] | <.0001 |
| LBBB vs Non-LBBB | 2.294 [1.952, 2.696] | <.0001 | 2,919 | 1.911 [1.593, 2.291] | <.0001 |
| LVEDV | 0.999 [0.998, 1.000] | 0.0024 | 3,739 | 0.998 [0.997, 0.999] | 0.0035 |
| LVEF | 0.996 [0.987, 1.005] | 0.4062 | 3,739 |  |  |
| NYHA I/II vs III/IV | 1.267 [1.111, 1.445] | 0.0004 | 3,728 |  |  |
| QRSd | 1.007 [1.005, 1.010] | <.0001 | 3,223 | 1.005 [1.001, 1.008] | 0.0107 |
| Renal Disease | 0.569 [0.475, 0.682] | <.0001 | 3,739 | 0.663 [0.532, 0.826] | 0.0002 |
| Female vs Male | 1.782 [1.530, 2.075] | <.0001 | 3,739 | 1.385 [1.144, 1.675] | 0.0008 |

### CRT response in NICM LBBB cohort

In a subgroup analysis of only patients with a NICM LBBB phenotype (1308 patients, 538 female), there was no significant difference in CRT response between males and females (73% vs 70%, p=0.25). A logistic regression model (Table 5) when applied to NICM LBBB patients showed that CRT response was significantly and independently associated with QRSd/LVEDV but not with sex. Normalised QRS was significantly different between females, who had median value of 0.919 (0.734-1.127) ms/ml, and males who had median value of 0.708 (0.585-0.886) ms/ml (p<0.001). For female patients with a QRSd/LVEDV of >0.915ms/ml (the median value), the CRT response rate was 78%, compared to 68% in patients where the QRSd/LVEDV was below the median value (p=0.012, Figure 1). CRT response as a function of QRSd/LVEDV for male patients is shown in Supplemental Figure 2.

**Table 5.** Logistic regression results (non-ischaemic left bundle branch block population)

| Parameters | Univariate |  |  | Multivariate (1295 patients) |  |
| --- | --- | --- | --- | --- | --- |
|  | Parameter Estimate<br>[95% CI] | p-value | Sample<br>Size | Parameter Estimate<br>[95% CI] | p-value |
| Age | 1.002 [0.991, 1.013] | 0.7481 | 1308 | 1.000 [0.988, 1.012] | 0.9983 |
| AF vs No AF | 0.651 [0.449, 0.945] | 0.0240 | 1308 | 0.627 [0.430, 0.913] | 0.0150 |
| COPD Yes vs No | 1.400 [0.908, 2.158] | 0.1275 | 1308 |  | . |
| Diabetes Yes vs No | 0.759 [0.585, 0.985] | 0.0378 | 1308 | 0.751 [0.577, 0.978] | 0.0333 |
| Hypercholesterolemia | 0.922 [0.715, 1.187] | 0.5278 | 1308 |  | . |
| Hypertension Yes vs No | 0.994 [0.780, 1.265] | 0.9590 | 1308 |  | . |
| LVEDV | 0.998 [0.997, 0.999] | 0.0041 | 1308 |  | . |
| LVEF | 1.009 [0.991, 1.026] | 0.3307 | 1308 |  | . |
| NYHA I/II vs NYHA III/IV | 1.115 [0.876, 1.419] | 0.3751 | 1304 |  | . |
| QRSd | 1.010 [1.004, 1.016] | 0.0022 | 1295 |  | . |
| QRSd/LVEDV | 1.948 [1.267, 2.996] | 0.0024 | 1295 | 1.984 [1.246, 3.157] | 0.0039 |
| Renal Disease Yes vs No | 0.616 [0.417, 0.909] | 0.0147 | 1308 |  | . |
| Female vs Male | 1.154 [0.903, 1.475] | 0.2509 | 1308 | 0.987 [0.758, 1.285] | 0.9202 |

**Figure 1:**
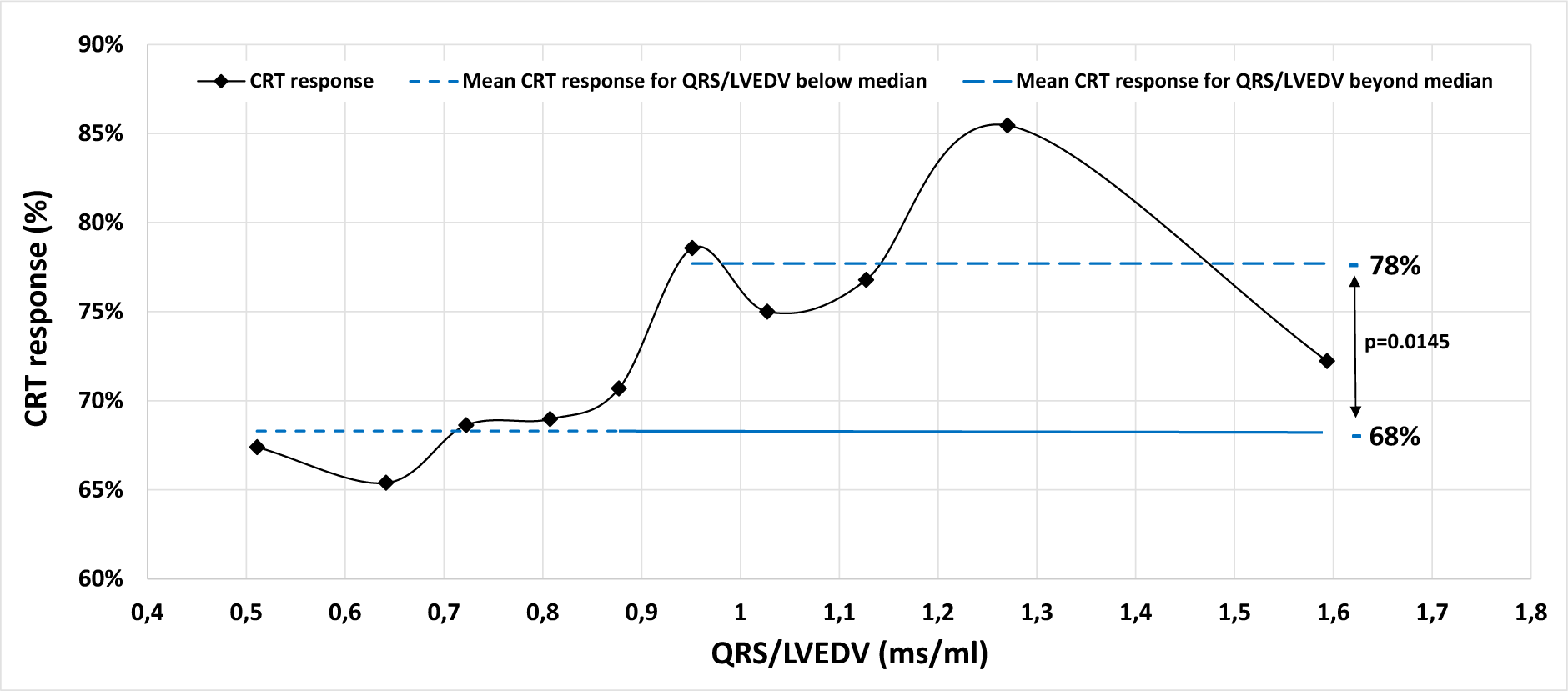
CRT response as function of QRSd/LVEDV in NICM LBBB female patients. QRSd/LVEDV was treated as a continuous variable and divided into 10 deciles.

### Analysis of sex-impact as a function of QRS in NICM LBBB cohort

In a further analysis of CRT response as a function of QRSd in patients with NICM and LBBB *and* the female CRT response rate was statistically superior to that of males (69.7% vs. 56.9%, p = 0.015) at QRSd < 150ms (n=315, with 54 patients displaying QRSd < 130ms) see Figure 2). The predicted probability of CRT response, estimated by the logistic regression model, shows that CRT response improves diminishing LVEDV and increasing QRSd/LVEDV and that at QRSd <150ms female patients have higher CRT response vs. male patients (Figure 3). A QRSd/LVEDV of 1.54 in patients with NICM, LBBB, and QRSd < 150ms [AUC: 0.62, 95% CI: 0.56-0.69; p=0.0002] was the optimal cut point predicting at least 80% CRT response, based on ROC analysis using the point on the curve closer to the upper left corner (Supplement Figure 3). Instead, when considering the entire spectrum of QRS durations, no difference in the predicted probability of CRT response was observed between the genders (Supplement Figure 4).

**Figure 2.**
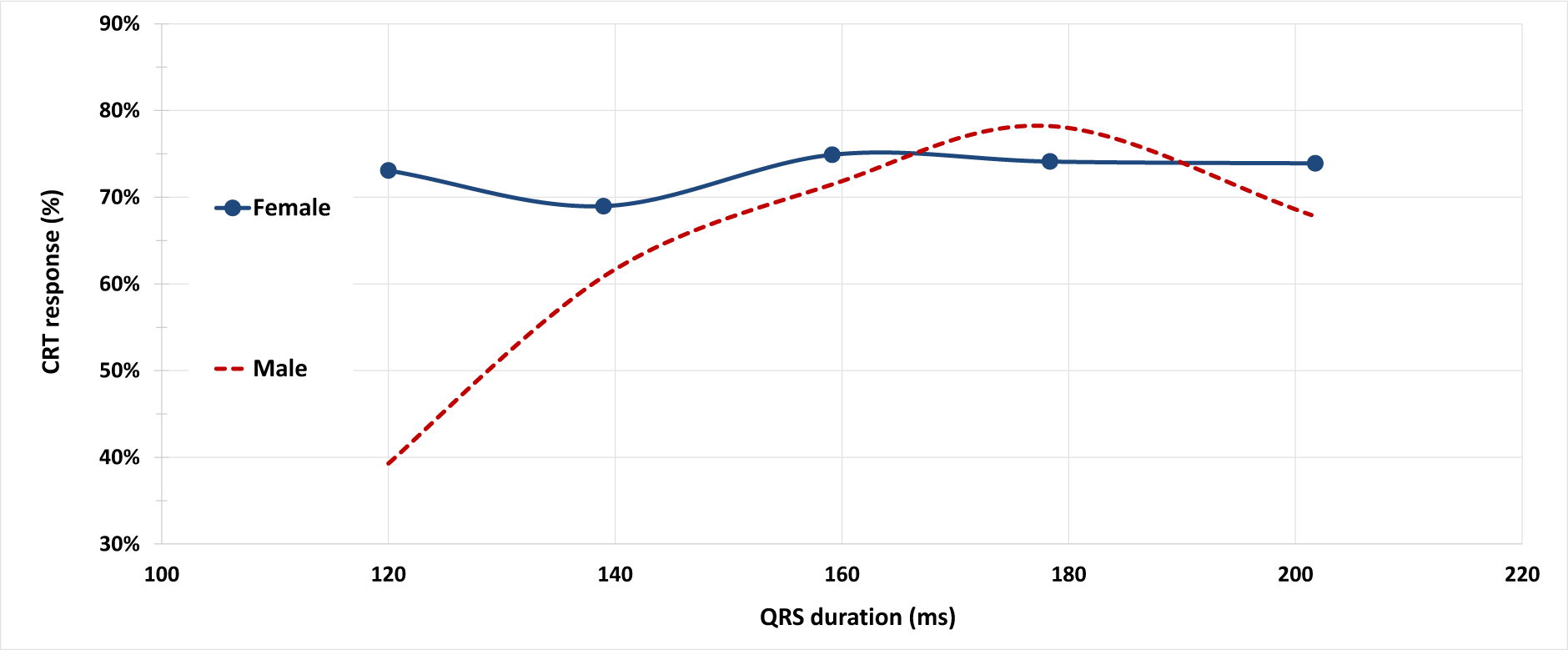
CRT response in patients with NICM and LBBB as a function of QRSd. QRSd was treated as a continuous variable, divided in 5 strata, as follows: strata 1 - QRSd<130ms; strata 2 - QRSd≥130 and <150 ms; strata 3 patients with QRSd≥150 ms and <170 ms, strata 4 patients with QRSd≥170 ms and <190 ms, strata 5 patients with QRSd ≥ 190ms.

**Figure 3.**
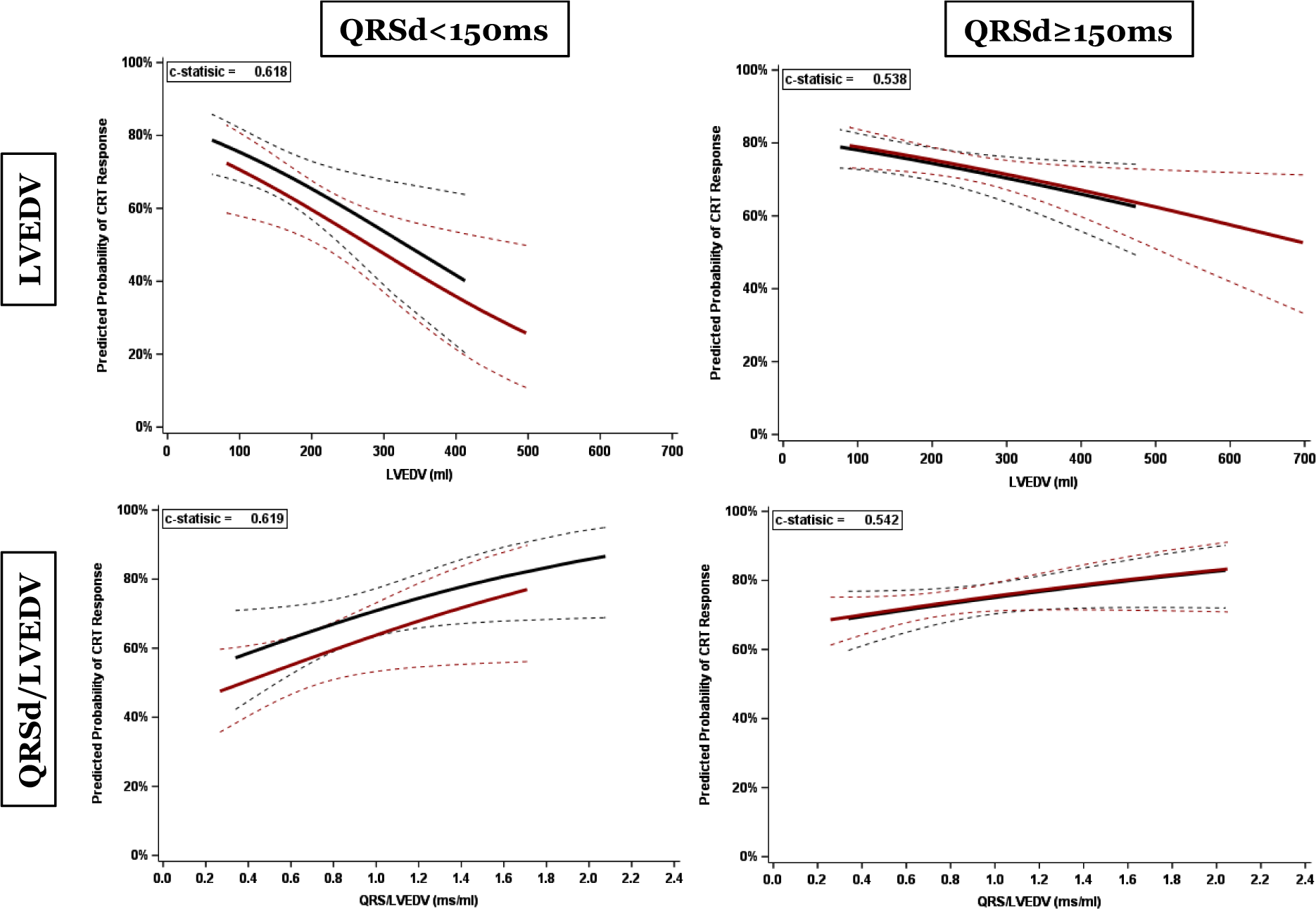
Parametric estimates with corresponding 95% confidence intervals showing predicted CRT response as function of LVEDV (top 2 panels) and normalised QRSd (bottom 2 panels). The left panels display data for patients with QRSd <150ms, and the right panels for those with QRSd >150ms. Blue (with 95% CI): female patients, red (with 95% CI): male patients.

Further parametric estimates are displayed in Figure 4, showing that the cohort with the highest predicted CRT response are female patients with an LVEDV < the median value, “smaller” heart size. The difference between male and female patents is not significant at larger heart sizes.

**Figure 4.**
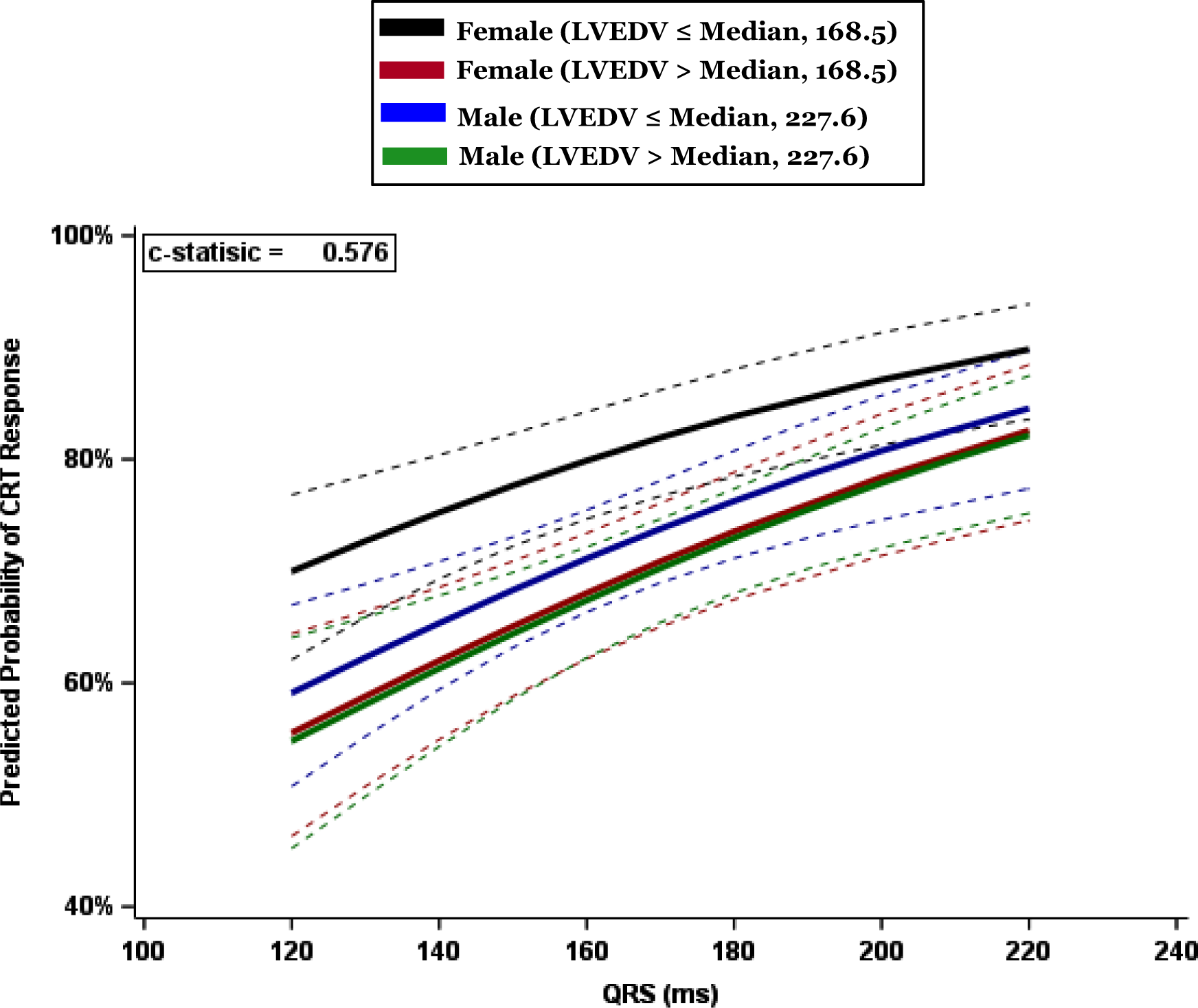
Parametric estimates with corresponding 95% confidence interval showing predicted CRT response as function of QRSd in 4 subgroups: females with heart size above and below median LVEDV of 168.5ml, and males with heart size above and below median LVEDV of 227.6ml.

Females with smaller heart size had a significantly higher CRT response than all three other cohorts (Supplement Table 1): females with a larger heart size (OR 1.87, 95% CI 1.26-2.77); males with a small heart size (OR 1.61 95% CI 1.11-2.33) and males with a large heart size (OR 1.92, 95% CI 1.31-2.82).

## Discussion

We describe here a post-hoc analysis from the prospective multicenter MORE-CRT MPP, a large trial of 3739 participants, all with complete echocardiographic data available at baseline and 6 months. This large sample size allowed associations between CRT reverse remodelling response and characteristics such as sex to be reliably determined, as well as provided the opportunity to analyze normalised QRS, a baseline variable which is difficult to examine without complete imaging datasets. Importantly, we report results from the largest dataset of female CRT patients ever evaluated so far.(14) Our findings were as follows:

1. CRT response was superior in females in the *overall cohort*.
2. In the *NICM LBBB cohort*, female sex was not an independent response predictor.
3. In the *NICM LBBB cohort,* increasing normalised QRSd is a significant independent response predictor.
4. In the *NICM LBBB cohort*, CRT response was superior in females at QRSd < 150 ms and the association between normalised QRSd and response is strongest at QRSd <150ms.

### Female sex and CRT response

In the MORE-CRT MPP population, CRT response was overall superior amongst female patients, supporting data from previous studies.(7–12) Our primary logistic regression analysis identified the female sex as a significant positive CRT response modifier, as well as the absence of AF, NICM, LBBB, and the absence of renal disease. Interestingly, however, when a subgroup analysis of 1308 patients with NICM and LBBB was performed to control for the effects of cardiomyopathy phenotype and QRS morphology, there was no significant difference in CRT response between female and male patients.

There is conflicting evidence for whether sex is truly an independent modifier of response or whether it is related to confounding variables. Arshad et al(24) suggested from a post-hoc analysis of the MADIT-CRT trial that sex discrepancy was related to a higher proportion of NICM LBBB patients in females, factors that are well-established as being beneficial in CRT response. Whilst previous studies have reported that the female sex may be an independent CRT response modifier, it is important to note the limitations of logistic regression analyses. Use of logistic regression to examine this association may be limited by several factors, such as: small female sample size in CRT trials; the assumption of linearity between female sex and CRT response; and the assumption of average or low multicollinearity between dependent and independent variables, such as sex and LVEDV.(25) Our analysis of a large subgroup of NICM LBBB patients is consistent with the theory that the female sex is not *independently* associated with CRT response, but rather, is related to confounding from positive response modifiers, such as NICM and LBBB.

### Sex-specific impact of QRS, heart size and CRT response

Our analysis has demonstrated that the sex-specific divergence in CRT response is most pronounced at shorter QRSd. This supports two previous secondary analyses from the MADIT-CRT study which reported that amongst patients with QRSd <150ms, only females derived benefit from CRT compared to ICD.(26,27) The reasons for this may lie in the discrepancy between male and female heart size, and thus, “relative dyssynchrony”, measured by normalised QRSd. Indeed we demonstrate that in the subgroup of NICM LBBB patients, QRSd/LVEDV is a significant predictor of CRT response. This association is strongest at narrower QRSd and smaller heart sizes. We suggest therefore that the sex-specific differences in CRT response in this subpopulation are a result of increased relative dyssynchrony in the females with narrow QRSd, but reduced heart size compared to their male counterparts, and thus, an increased QRSd/LVEDV ratio. This supports previous smaller studies which have suggested that normalized QRSd may be an appropriate target to guide CRT implantation,(19–21) and mechanistic studies which have reported that echo-derived dyssynchrony was more predictive of CRT response than absolute QRSd, and may be beneficial in patients with a narrow QRSd.(28,29) Furthermore, a post-hoc analysis of the ECHO-CRT trial, a study which reported futility in patients with a narrow QRSd implanted with CRT, reported that: 1) males formed the majority of patients and they drove the negative outcome, while CRT-ON vs. CRT-OFF comparison was neutral in females; 2) the higher risk of negative outcome was concentrated among those with larger LV dimensions and 3) CRT, compared with the control group, induced significant LV reverse remodelling in patients with large normalized QRSd/LVEDV (>1.3 ms/ml).(30)

It should be noted that our results indicate that the strength of the correlation between sex and CRT response appears to be attenuated at very high QRSd. We theorize that this is because at these high levels of relative dyssynchrony, CRT response rates are high in both sexes (approximately 70-75%). Therefore, despite increased QRSd/LVEDV in female patients at these levels, alternative issues are likely to be driving non-response, thus diminishing the effect of sex on CRT efficacy at the high extremes of QRSd.

### Clinical implications

This study demonstrates that heart size is an important factor in driving sex-specific CRT response, in view of increased normalised QRSd in female patients, with the benefit predominant at narrower QRS durations. These associations were observed through a sub-group analysis of the NICM LBBB cohort which mitigated noise from confounding variables.

Within an NICM LBBB population, selecting CRT recipients based on absolute QRSd dichotomization may exclude certain female patients who could benefit from treatment based on a high degree of relative dyssynchrony due to small heart size. Current European and US guidelines define QRSd cut-offs of 130ms and 120ms respectively, as the target criteria for CRT implantation.(1,31,32) These cut-offs are defined from meta-analyses of studies with few female participants, which may hide a possible beneficial effect of CRT in female patients with narrower QRSd.(33) This study opens the debate on whether the use of normalized QRSd should be integrated into routine clinical practice to identify these patients. Alternatively, sex-specific QRSd cut-offs may be considered as a practical surrogate accounting for the significant disparities in normalised QRSd.

### Limitations

This study represents a retrospective analysis of the observational MORE-CRT MPP trial. As such, we cannot exclude inherent limitations associated with observational studies, such as selection bias. Nevertheless, the study has important strengths: 1) data collection was prospective; 2) analyses objectives were prespecified; 3) monitoring with strict source data verification activities; 4) echocardiographic evaluations by an echo core lab and 5) the large sample size allowed us to control for confounding patient characteristics, such as ischemic vs non-ischemic cardiomyopathy or LBBB vs. non-LBBB QRS morphology. Overall, we are confident that the study data provides a fair description of current CRT application and outcomes, however, our conclusions should be interpreted as hypothesis generating.

## Data Availability

Data can be requested from the study sponsor.

## Acknowledgment

The authors thank all investigators, the staff, and the participants of the MORE CRT MPP trial for their valuable contributions. A full list of participating principal investigators and institutions can be found in Supplemental Table 2.

